# A Modified Version of the Function In Sitting Test (mFIST): Development and initial Reliability of the mFIST in Outpatient Clinic in Veterans with Spinal Cord Injury

**DOI:** 10.1101/2025.08.29.25334480

**Authors:** Nichole Vasquez, Valerie Chavez, Perri Kraus, Maureen Jennings, Joayn Troung, Tamara Alexander, Maya N. Hatch

## Abstract

**Introduction:** Trunk control and seated balance are important for various activities of daily living in patients with spinal cord injuries (SCI). A clinically appropriate and reliable gold standard seated balance outcome measure for SCI is lacking. The Function in Sitting Test (FIST) has become popular in stroke and in-patient clinics for tracking seated balance. A modified version of the Function in Sitting Test for SCI patients may prove similarly useful for measuring and tracking seated balance in the clinic.

**Objective:** The primary objective of this study was to modify the FIST for the SCI population (mFIST) and assess initial reliability of the measure in a busy outpatient, real-world, clinical setting. The secondary objective was to determine trends in scoring differences if reliability was below excellent.

**Design:** Prospective reliability study. The mFIST was administered to Veterans with SCI twice within 2 weeks by the same evaluator (test re-test), while being video-recorded for subsequent scoring.

**Setting:** Busy Outpatient Clinic in Veteran Affairs Healthcare System.

**Participants:** A total of 42 Veterans with SCI (mean age = 62, 91% male), between C4-L1 and with an American Spinal Injury Association Impairment Scale grade of A to D, participated in this study during routine outpatient clinic. Participants had to be primary wheelchair users and able to sit unsupported for at least 1 min.

**Interventions:** Not Applicable.

**Main Outcome Measures:** Test re-test, intra-rater and inter-rater reliability using intra-class correlation coefficients were determined. Mean rating differences were calculated for individual mFIST items, total scores across all subjects, and when separated by SCI injury level grouping to investigate reliability trends.

**Results:** Test re-test and intra-rater reliability for the mFIST for the entire cohort were good with ICCs of 0.88 (CI 0.77 – 0.93) and 0.89 (CI 0.51-0.97), respectively. Inter-rater reliability was excellent with ICC = 0.92 (CI 0.85 – 0.96) across the entire cohort, however reliability dropped (0.78; p= 0.02) for certain SCI subgroups. The largest differences in scoring across reliability testing were seen during dynamic tasks in those with cervical injuries.

**Conclusion:** The mFIST displays good to excellent reliability during routine use in a busy outpatient clinic. Inconsistencies in scoring of some of the dynamic items, however, indicate that the measure might need further wording and/or scoring refinements before being widely distributed. Determining the validity and sensitivity of the mFIST is also needed.

## Introduction

In the United States, as many as 393,913 people are currently living with a spinal cord injury (SCI).^1^ Damage from a spinal cord injury results in varying degrees of neurologic impairments and sensorimotor damage^2,3^ that can result in trunk instability and seated balance impairments. Prior studies have shown that impaired sitting balance in patients with SCI affects a variety of daily activities such as dressing, reaching for items, positional changes (including wheelchair propulsion), transfers, and even eating.^4–7^ Ability to control seated balance is thus important for daily life and independence.

There is currently no gold standard to measure seated balance/ trunk control in individuals with SCI. This is to the detriment of the field as a wide range of new interventions and therapies showing potential for improving trunk control are entering the SCI space.^8–10^ Traditional outcomes used in SCI for balance are cost and space prohibitive (i.e. posturography)^11,12^, are primarily for those that can stand (i.e. berg balance scale)^13^, or are unidimensional (i.e. modified Functional Reach Test)^14^; resulting in poor clinical utility. The Function in Sitting Test (FIST), developed by Gorman et al^15^ has gained popularity as a clinical assessment of seated balance due to its ease of use in the clinic and comprehensive testing (i.e. static, dynamic and reactive balance). Although originally validated in stroke patients, the FIST has also been validated in patients with multiple sclerosis and adults in inpatient rehabilitation^16,17^, and has potential applicability for SCI.

Abou et al^18^ tested the FIST in a small cohort of non-ambulatory SCI subjects. The FIST was found to have excellent test re-test reliability but only satisfactory internal consistency, which they attributed to some of the items not being relatable to the SCI population, suggesting modifications would be needed. This study also lacked inter-rater reliability and heterogeneity of SCI participants, thereby limiting its generalizability.

Palermo et al^19^ modified the FIST (aka FIST-SCI) by removing the stroke-specific items making it more applicable to the SCI population. Initial data was collected from 38 SCI participants. Results indicated excellent inter-rater reliability, intra-rater reliability, and internal consistency. The FIST-SCI was also able to detect independent transfer ability with high sensitivity and specificity (92%). During the Palermo publication, our team was also modifying the FIST for the SCI population and amid reliability testing on a more generalizable SCI population. Our team’s modification approach was slightly different than Palermo’s. Both of our versions included the original 14 items for testing. However, Palermo’s FIST-SCI altered the scoring scale (removing the verbal cueing and expanding the level of assistance), added bilateral performance of some tasks (instead of just the stronger side) and using the lower score, and based on our understanding loosened some requirements of movements to the fullest range of motion. In our modified version of the FIST, we elected to keep the same scoring scale as the original FIST (Palermo’s FIST-SCI altered the scoring system) and we maintained the strict movement criteria to receive the top score while providing some downgrading of scores if criteria was not met. In this study, we also changed the initial experimental design. Both Abou et al^18^ and Palermo et al^19^ performed reliability testing as a side portion of another research study and not during routine clinics. Limited numbers of raters for reliability testing were also used; 1 rater in Abou and 2 in Palermo. Raters in Palermo et al assisted with the creation of the FIST-SCI, which may bias reliability to higher agreement levels compared to what may occur during widespread deployment and use. As a rule of thumb, previous work has suggested reliability studies should try to conduct testing on 30+ heterogenous sample subjects with at least 3 different raters.^20^ This allows greater chances of capturing differing levels of variability at the subject and rater levels. Psychometric testing of a new measure should also be performed in an environment that is as close to reality as possible, representing real world application. Most rehabilitation and/or SCI clinics will involve several physical therapists /raters at any given time, with varying levels of expertise.

The primary goals of this study were to: (1) briefly describe our modified version of the FIST (mFIST) for SCI; and (2) perform test re-test, intra-rater and inter-rater reliability on the mFIST in a busy outpatient clinical setting and identify the items with the most variability in scoring if correlation coefficients were not excellent. Our experimental design is distinct in that: we tested patients during existing clinical appointments with typical space and time constraints; at least 3 raters (up to 6) were used for reliability testing with majority being naïve to the measure and the modification process; and testing was performed on a heterogenous SCI cohort. The secondary goal of this study was to investigate if patterns of rater differences exist across mFIST items and/or certain SCI-subgroups.

## Methods

### Modifications to FIST

The original FIST is composed of 14 items that include 4 static or sitting tasks, 7 dynamic, and 3 reactive balance tasks, with a rating scale from 0 (complete assistance) to 4 (independent) for each item.^15^ Total scores can range from 0 - 56. Mean FIST scores across majority of the SCI subjects in Abou et al^18^ including cervical injuries, were all around the top 3^rd^ of the scale. This leaves little room for detecting significant change over time or when describing different levels of seated balance dysfunction. It can also lead to ceiling effects, thereby limiting its clinical utility for SCI. The authors also mentioned that several tasks were considered “not challenging enough”, or not relevant to those with SCI.

Our goal was to keep the number of testing items and scoring scale similar to the FIST, owing to its existing success in other conditions, but updated items to make them relevant to individuals with SCI and improve applicability by increasing the range of potential scores.

Two rehabilitation experts (with SCI experience) and one researcher participated in the modification process. Phase 1 included modifications to address the physical limitations of those with SCI which included: changing the item for retrieval to an empty water bottle (instead of a small tape measure that would require fine motor skill), allowing a one arm leg lift for those without hip flexors, and loosening requirements for arm abduction or flexion to shoulder level during leaning tasks.^15^ Phase 2 occurred after beta testing on a few subjects after which modifications to wording, updates to item testing order, and other changes to limit compensations and ensure proper technique (use of trunk musculature) to obtain a full score for: forward reach, lateral reach, and picking up object from behind. Reactive and static sitting tasks were kept as originally scored and described, apart from sitting while lifting the thigh. Please refer to Supplemental Table 1 and Supplemental Table 2 for comparison of mFIST changes and final mFIST scoring sheet. Photographic images of demonstrating each of the 14 testing items are available upon request.

### In-Clinic Recruitment

Of the ~363,000 individuals in the United States are currently living with an SCI 78% are males. The Veteran’s Health Administration (VHA) provides comprehensive care to ~42,000 Veterans with SCI, or 17,500 new cases each year,^21^ making it the single largest network of providers for individuals with SCI. Services for SCI include comprehensive annual evaluations, and a variety of outpatient clinics to address impairments in function and mobility, including wheelchair clinics. Veterans with SCI were approached for study participation during their routine outpatient clinic appointments at the Long Beach VA Medical Center in the Spinal Cord Injury /Disorder (SCI/D) center by one of the therapists/team members. Study inclusion was purposefully kept as broad as possible to attract a variety of subjects. The following inclusion criteria were used to identify participants: ≥ 18 years old; SCI level between C4 - L2 with ASIA Impairment Scale (AIS) grade A through D; >12 months post injury; primary wheelchair user (defined as > 50% time in wheelchair); ability to comprehend and follow study procedures; ability to sit unsupported and unassisted for brief periods; and at least 2 scheduled appointment across a 2 week time period. Exclusion criteria included: primary neurological injury other than SCI; documented balance disorders (i.e. Parkinson’s, brain injury/trauma, inner ear disorders, or hemiparesis); untreated spasticity; current pressure ulcer in the trunk/pelvic area or upper thighs; inability to achieve short-sitting position (90° flexion at hip and knee); ventilator dependent; and unstable angina and/or presence of orthostatic hypotension. International Standards for Neurological Classification of Spinal Cord Injury (ISNCSCI) exam must have been performed within the last 6 months at the center and documented in the VHA electronic medical record.

This protocol was approved by the Long Beach VA institutional review board and written informed consent was obtained for all study participants prior to data collection. The first visit was performed either on the same day as consenting (often the case for those in Annual Evaluation or wheelchair clinics) or on subjects next clinic appointment.

### Clinical and demographic information

Basic demographic and SCI clinical information was collected from participants using the NIH Common Dataset (F1737 and F1790, respectively).^22^ Demographic information included, age, sex, race, ethnicity, approximate weight, marital status and occupation. All other categories in the F1737 were collected but not reported. Clinical information included injury duration, type, and etiology for both non-traumatic and traumatic injuries. Neurological injury level and completeness were pulled from the subjects recent ISNCSCI exam from the electronic records system.

### Test Re-Test, Intra- and Inter-rater Reliability Procedures

This study was designed to test all 3 types of reliability on the outcome measurement of interest (mFIST: Figure 1). Clinical evaluators were provided with a brief introduction to the mFIST and were instructed to read and perform the mFIST based on the instructions on the scoring sheet. They were also instructed to use the provided wording for each item, as much as possible. Some evaluators had prior experience with the original FIST, and others had limited experience. Reliability calculations for the entire cohort and across SCI sub-groups were performed. SCI sub-groups included: cervical (C2-C7), high thoracic (T1-T6), and low thoracic+ (T7 - L2). Division of thoracic injuries at the T6 level was determined based on innervation of the trunk muscles: with majority of the trunk muscle innervation at T7 and below.^23–25^

**Figure 1:**
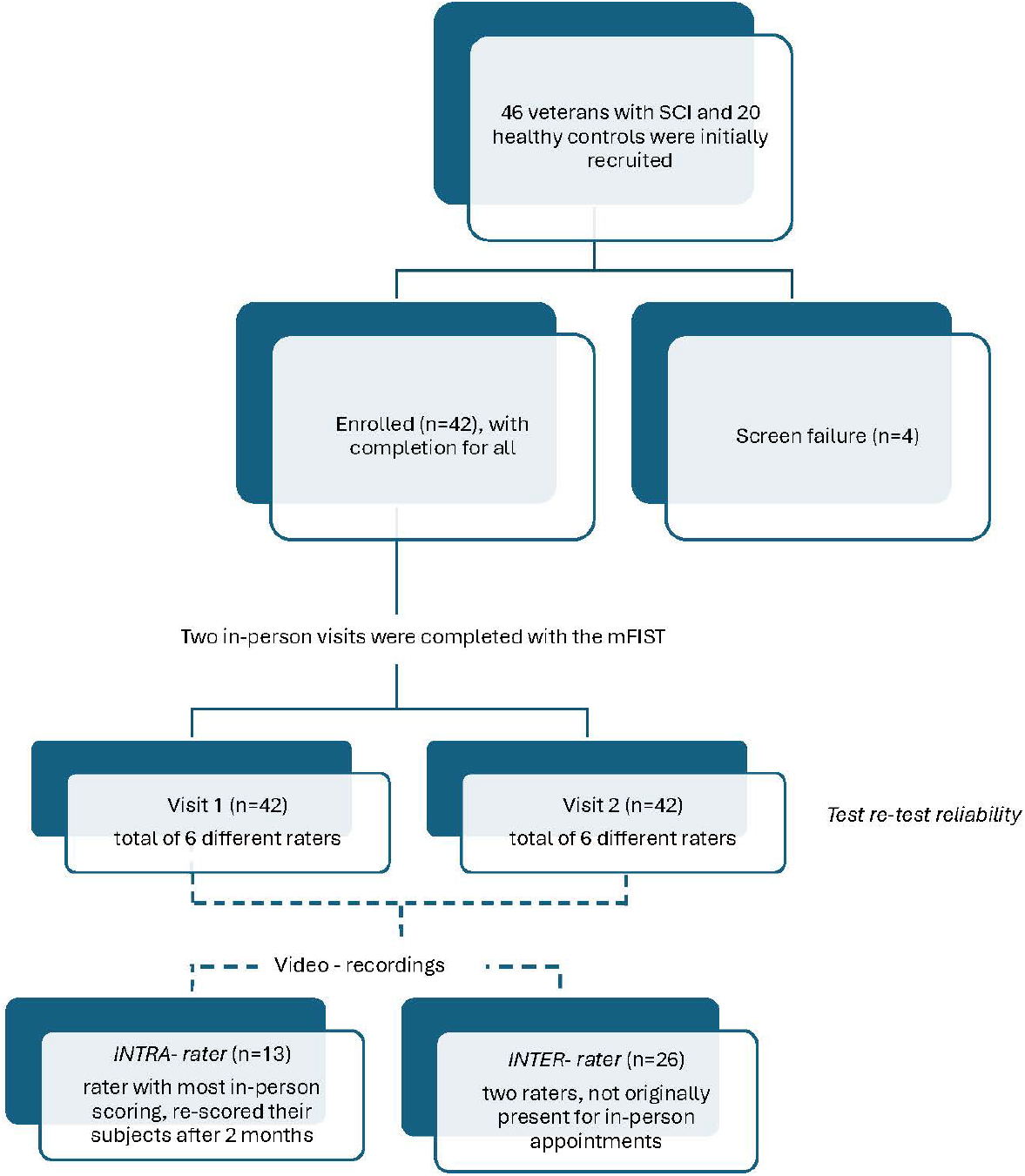
Study flowchart. Numbers of subjects recruited and enrolled in the study. All subjects enrolled completed two visits separated by at least 24 hours and within 14 days. Subject visits were video recorded for intra- and inter-rater analyses.

#### Test Re-Test

The mFIST was administered twice in-person to participants with SCI by the same rater (Figure 1), while also being video recorded for both sessions. All sessions were performed at the beginning of the clinical appointment. The two visits were separated by at least 24 hours but within 14 days. All SCI subjects were at least 1 year post injury and thus presumed to be functionally stable. Four primary raters were utilized for majority of the in-person scoring, with two additional raters being used sparingly (< 2% of subjects tested) due to their involvement in creation of the mFIST (MH and MJ). This methodology targeted reliability from a variety of raters across a wide range of individuals with SCI. All videos were recorded using the same VA-issued iPAD on a tripod where videos were kept behind the VA firewall until they were transferred to a study specific (protected) folder for storage. Video angles were consistent across subjects and visits and included front and side views, depending on the testing item.

#### Intra-rater

Understanding that individuals with SCI may have inherent variability in performance across the two in-person visits irrespective of the rater, a separate method for intra-rater reliability was performed. The rater with the highest number of in-person subjects (n=13 subjects) re-scored individuals by video at least 2 months after all their subjects had completed participation (Figure 1). Two months allowed enough time to forget the details of any one subject’s performance. Re-scoring the exact performance of the subject in this manner was deemed a true within-rater assessment. Subjects were re-scored in random order.

#### Inter-rater

Two raters (MH, VC), who were not originally present at the in-person appointments, independently scored half of the subjects (n=26) by video. The same videos were used for both raters (Figure 1). Scores were compared to the original, in-person, ratings across all subjects. Sample subjects were chosen by the coordinator who attempted to balance the distribution of subjects across SCI injury sub-groups.

#### Rating differences

Differences between individual ratings across mFIST items were investigated to identify patterns of rating differences. Differences for test retest were calculated by: |visit rating_2_ – visit rating_1_|, where rating_1,,2_ were in-person ratings by the same evaluator.

Differences for intra-rater were: |rating_2_ – rating_1_|, where rating_2_ were second rating by video and rating_1_ was the first rating inperson (base rater). Differences for inter-rater were: |rating_2,3_ – rating_1_|, where rating_2,3_ were second or third ratings by video and rating_1_ was the first rating in-person (base rater). Rating differences were calculated for mFIST total scores across all subjects, and when separated by SCI injury level grouping. Item differences were also grouped into 1 of 3 testing domains (static, dynamic, reactive) and summed to investigate whether types of testing domains were more prone to rating differences. Static tasks included sitting still with eyes open & closed; sitting while rotating head; and sitting with 1 leg lift. Dynamic tasks included picking up objects from behind and floor; scooting in all directions (behind, side and forward); and lateral and forward reaching. Reactive tasks included anterior, posterior and lateral nudges.

### Statistical Analyses

All analyses were conducted using Stata version 16.2 software (StataCorp, College Station, TX). Basic statistics using frequencies, mean and standard deviations were used to report demographic information, SCI characteristics, and all rating differences.

Reliability was determined using Intra Class Correlation, absolute agreement options: A one-way random effects model with *k* raters was chosen for test re-test reliability because each subject was rated by a different rater. Two-way random effects models were chosen for both intra-rater and inter-reliability, with 1 or *k* raters respectively. These are equivalent to Shrout and Fleiss conventions^26^: ICC (1,*k*), ICC (2,1) and ICC (2,*k*) for test re-test, intra-rater and inter-rater reliability. Averages and corresponding confidence intervals were used. Correlation coefficients were performed on total mFIST scores for the entire cohort and on SCI sub-groups (cervical, high thoracic, and low thoracic) for test re-test and inter-rater. Sub-group analyses were not performed for intra-rater due to the small numbers of subjects in each group (<5). A p-value of 0.01or less was used to determine significance.

## Results

### Cohort Characteristics

A total of 46 Veterans with SCI were approached during routine clinics. Four Veterans with SCI were excluded due to screen failure and one participant was unable to complete the second visit, leaving a total of 42 subjects that completed study procedures (Figure 1).

Our study cohort primarily consisted of Non-Hispanic White males (91%), mean age of 62+ 12.7 yrs.

Majority lived in a big city (66.6%) and were retired with disability (64.3%; Table 1A). Majority had a traumatic SCI (76.2%), with even amounts of cervical (45.2%), and thoracic (54.8%) injuries (Table 1B). Only 1 individual with a lumbar injury (L1) was enrolled and was grouped with low thoracic grouping.

**Table 1:** Cohort characteristics.

| | Total<br>% (n) | Average<br>(±SD, if appropriate) | Range<br>(if applicable) | |
| --- | --- | --- | --- | --- |
| **A. Demographics (n=42)** |  |  |  |  |
| Age (yrs) | | 62±12.7 | 27-77 | |
| Reported weight (lbs) | | 188.2±49.5 | 107-380 | |
| Gender |  |  |  |  |
| Female | 7.1 (3) | - | - |  |
| Male | 90.5 (38) | - | - |  |
| Missing / not reported | 2.4 (1) | - | - |  |
| Race / ethnicity |  |  |  |  |
| Non-Hisp White | 54.8 (23) | -- | - |  |
| Hispanic White | 21.4 (9) | - | - |  |
| Black | 16.7 (7) | - | - |  |
| Missing / not reported | 7.1 (3) | - | - |  |
| Current occupation |  |  |  |  |
| Paid | 2.4 (1) | - | - |  |
| Retirement disability | 64.3 (27) | - | - |  |
| Retired | 11.9 (5) | - | - |  |
| Unemployed | 7.1 (3) | - | - |  |
| Missing / not reported | 4.8 (2) |  |  |  |
| **B. Spinal cord injury characteristics** |  |  |  |  |
| Injury duration (yrs) | | 22.7 ± 18.6 | 1.1 – 57.5 | |
| SCI level |  |  |  |  |
| Cervical | 45.2 (19) |  |  |  |
| High thoracic | 26.2 (11) |  |  |  |
| Low thoracic | 28.6 (12) |  |  |  |
| Injury type |  |  |  |  |
| Non-traumatic | 21.4 (9) |  |  |  |
| Traumatic | 76.2 (32) |  |  |  |
| Missing | 5.3 (1) |  |  |  |
|  | Cervical<br>% (n) | High thoracic<br>% (n) | Low thoracic<br>% (n) | Total<br>% (n) |
| AIS – grades |  |  |  |  |
| Complete (AIS A and B) | 21.1 (4) | 63.6 (7) | 66.7 (8) | 45.2 (19) |
| Incomplete (AIS C and D) | 78.2 (15) | 36.4 (4) | 33.3 (4) | 54.8 (23) |

### mFIST scores

Overall mean mFIST total scores (Figure 2) were 44.6 + 9.3 and 45.9 + 8.5 for visit 1 and 2, respectively. Mean total mFIST scores were fairly consistent across SCI injury groups: 45.1 + 10.2 and 47.9 + 6.9 for cervical; 44.2 + 8.4 and 43.9 + 8.2 for high thoracic; and 44.3 +10.0 and 44.8 + 12.2 for low thoracic for visit 1 and 2 respectively. Mean testing times were 14.0 + 7.4 min and 9.5 + 4.0 min for visit 1 and visit 2, respectively. Mean number of days between testing was 4 days.

**Figure 2:**
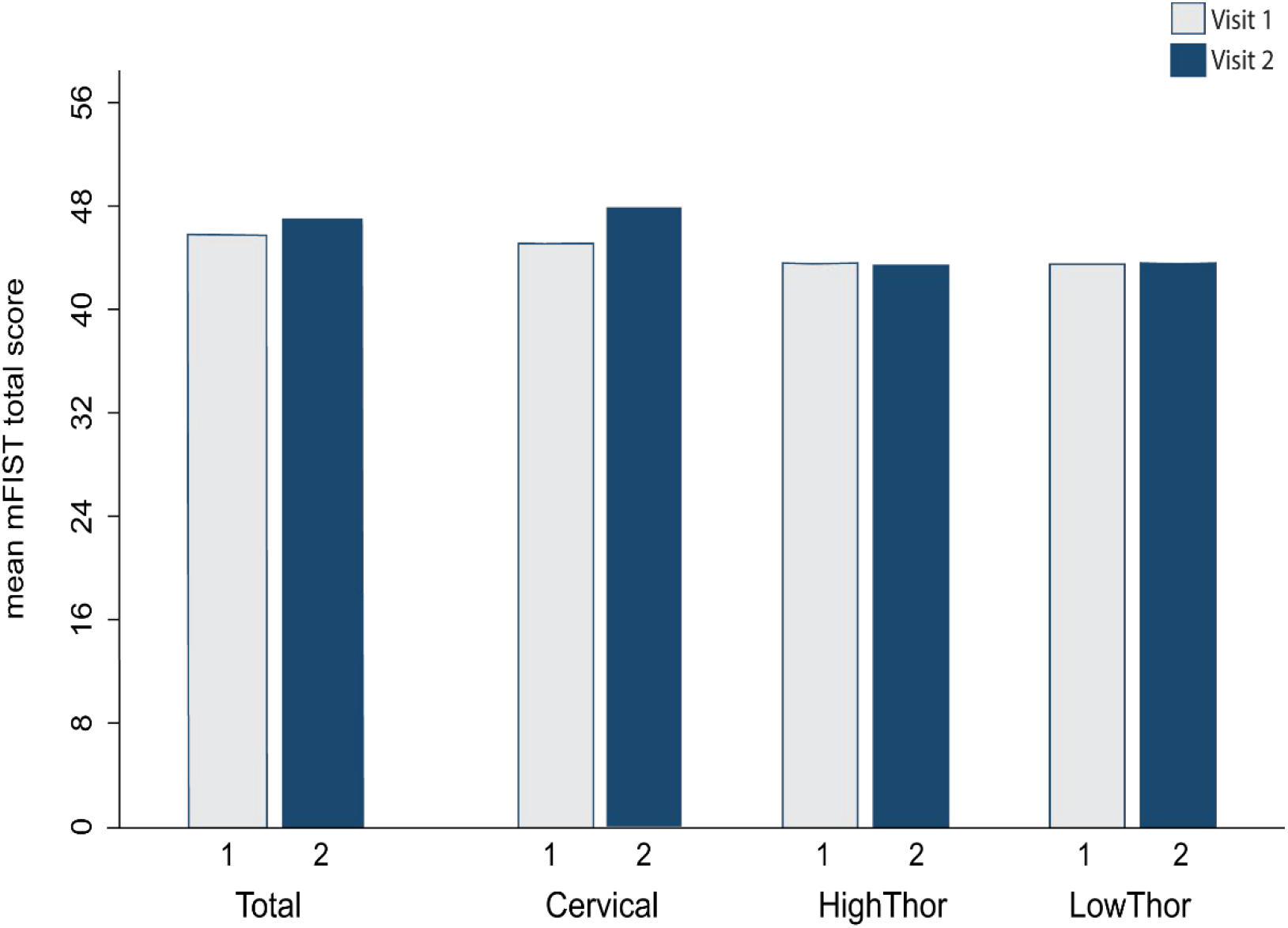
Mean mFIST total scores. Mean total scores for mFIST for visit 1 (grey) and visit 2 (blue) for the entire cohort (Total) and when broken down by SCI injury group. Cervical represents C4 - C7, High Thoracic T1 - T6 inclusive, and Low Thoracic for T7 and below. Max total score for mFIST is 56.

### Reliability and Scoring Differences

Test re-test reliability (Table 2) for mFISt total scores were considered good for the entire cohort (ICC = 0.88, p<0.001). However, when breaking the cohort into SCI sub-groups, reliability remained good for the cervical group (ICC = 0.78, p = 0.001), while mFIST total scores for high and low thoracic groups (ICC = 0.91 and 0.98, p<0.001, respectively) displayed excellent reliability (Table 3). Differences between total mFIST scores from visit 1 and 2 were higher in the cervical and high thoracic subjects (4.7 and 4.5, respectively) compared to lower thoracic (2.4; Figure 3A). When breaking up the items into testing domains, mean scores differed by 2.8 for cervical and high thoracic subjects for dynamic tasks, compared to 1.4 in low thoracic (Figure 3B).

**Table 2:**
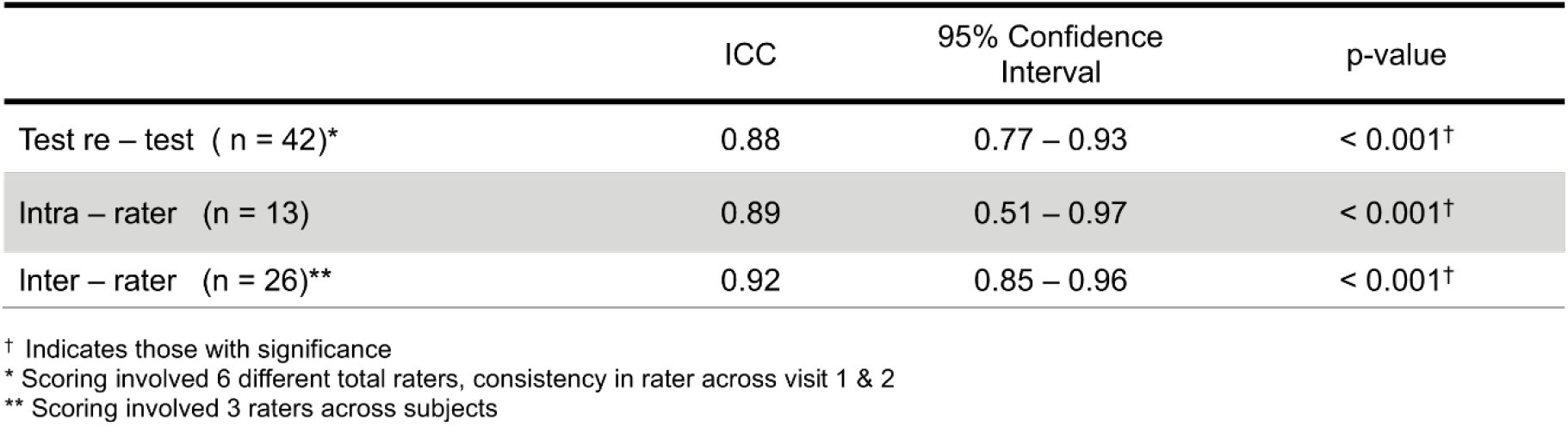
Reliability for mFIST total scores.

**Table 3:** mFIST reliability across SCI sub-groups.

|  | ICC | 95% Confidence Interval | p-value |
| --- | --- | --- | --- |
| Test re – test ( n = 42) |  |  |  |
| Cervical | 0.78 | 0.45 – 0.92 | < 0.01 |
| High thoracic | 0.91 | 0.73 – 0.97 | < 0.001† |
| Low thoracic | 0.98 | 0.89 – 0.99 | < 0.001† |
| Intra-rater<sup>‡</sup> | - | - | - |
| Inter – rater (n = 26) |  |  |  |
| Cervical | 0.90 | 0.75 – 0.97 | < 0.001† |
| High thoracic | 0.78 | 0.13 – 0.97 | 0.02 |
| Low thoracic | 0.98 | 0.94 – 0.99 | < 0.001† |

† Indicates those with significance
<sup>‡</sup> Sub-grouping could not be performed due to small sample sizes in each group ( $n \leq 5$ per group)

**Figure 3:**
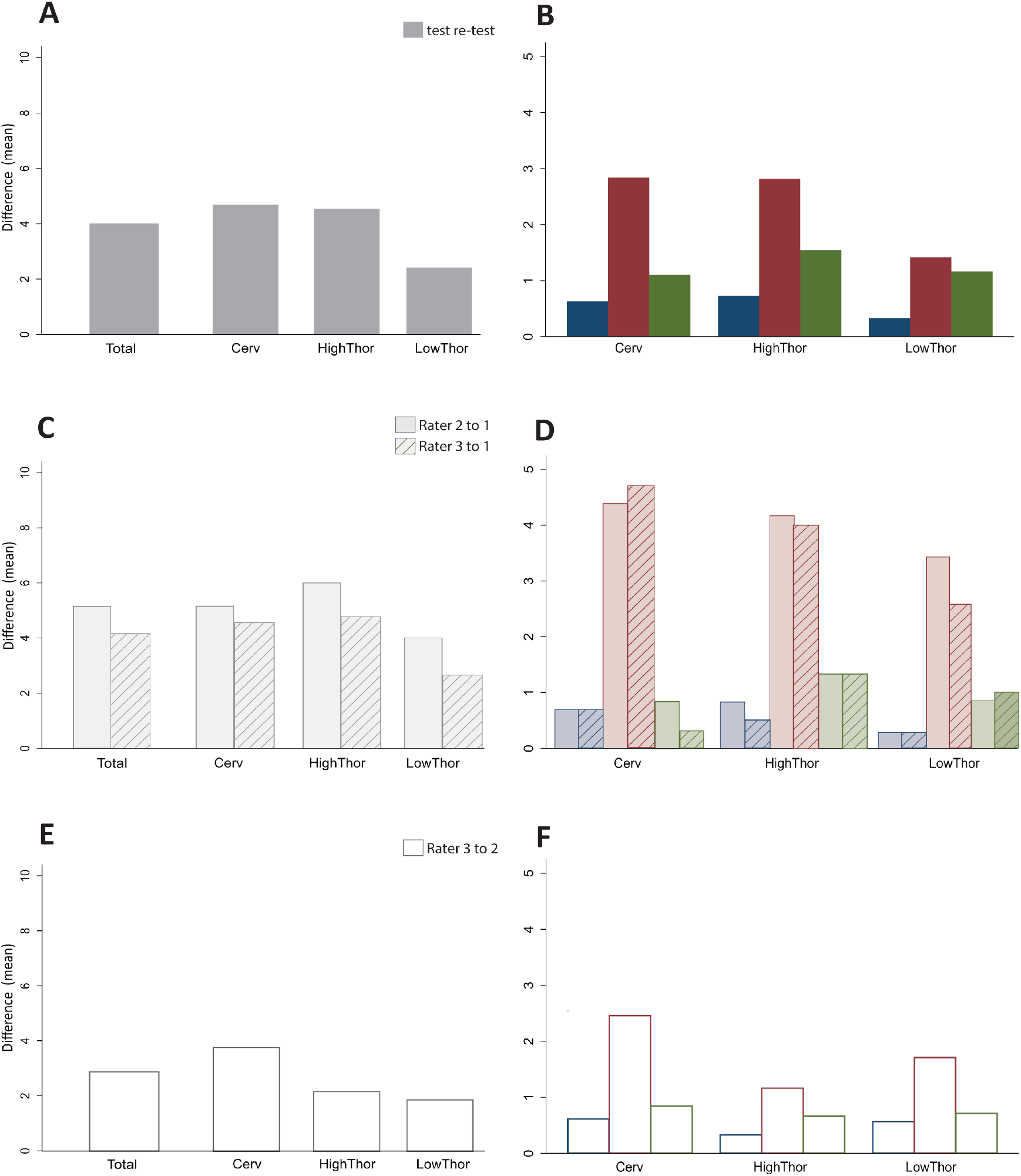
**(A-B)** Mean mFIST differences during in-person test re-test for total cohort, and when separating subjects by SCI sub groupings (Cervical, high thoracic, and low thoracic). **(C-D)** Mean mFIST differences during interrater for rater 2 (solid bars) and rater 3 (striped bars) when compared to the base rater (in-person rater). **(E-F)** Mean mFIST differences between rater 3 and rater 2. All blue colored (dark or light filled, striped and empty) represent static sitting items. All maroon colored (dark or light filled, striped and empty) represent dynamic items. All green colored (dark or light filled, striped and empty) represent reactive items. A, C, E represent total mFIST scores. B,D,F represent static, dynamic & reactive subtotals.

Intra-rater reliability was considered good for the entire cohort with similar coefficients to test re-test (ICC = 0.89, p< 0.001; Table 2). Differences between total mFIST scores were higher across dynamic tasks, (data not shown) compared to sitting and reactive tasks; keeping in trend with test re-test.

Inter-rater reliability (Table 2) was excellent for the entire cohort (ICC = 0.92, p< 0.001). When breaking subjects into SCI sub-groups, reliability was good (ICC = 0.78, p = 0.02) for high thoracic subjects, compared to cervical and low thoracic subjects which displayed excellent reliability (ICC = 0.90 and 0.98, p<0.001), respectively (Table 3). Inter-rater differences for mFIST scores were first performed by raters to the base rater. Differences between mean total mFIST scores were slightly higher for rater 2 (solid bars; 5.0), than rater 3 (stripe bars; 4.1) for the entire cohort (Figure 3C). There was a general trend for higher rating differences during dynamic tasks (solid and stripe pink bars) for both raters across cervical, high thoracic and low thoracic sub-groups (4.3, 4.2 & 3.4 for rater 2; and 4.7, 4.0 & 2.6 for rater 3), respectively, compared to sitting and reactive items (most < 1, lt blue and green; Figure 3D). When comparing rater 2 to 3 to each other, mean differences in total mFIST scores (Figure 3E) were lower overall (2.8) and across cervical (3.7), high thoracic (2.2.) and low thoracic (1.9) sub-groups, than when compared to the base rater (Figure 3C). There was a consistent trend for higher mean differences in dynamic tasks (open maroon bars; Figure 3F) across cervical (2.4), high thoracic (1.2), and low thoracic (1.7) sub-groups, albeit mean differences across the board were lower.

## Discussion

Clinical outcome assessments (COA) are critical components of clinical care and clinical trials: they are used to establish baseline function, detect changes over time, and determine the efficacy of a new treatment or intervention. With such a high level of importance, we must ensure that any new clinical outcome measure is properly examined before being released for wider use. Central to these requirements are studies investigating the real-world performance of a given clinical outcome measure.

The mFIST showed good test re-test and intra-rater reliability and excellent inter-rater reliability in our cohort of Veterans with SCI during routine clinical testing. These findings are slightly different than Abou et al who showed excellent test re-test for the traditional FIST on subjects with SCI. Although our test re-test reliability was near the clinical marker for excellent (set at 0.90), Abou et al’s was at the higher range of excellent (0.95). A notable difference between studies is that our cohort had much higher numbers of cervical injuries (n=19) than Abou’s whose was dominated by thoracic injuries (only 3 of the 26 were cervical). This is an important difference because when we divided our cohort into injury sub-groups, the cervical group displayed lower reliability (0.78) compared to both thoracic sub-groups (0.91 and 0.98). Majority of the subjects in the cervical group had incomplete injuries (AIS C or D) with a predominance of AIS D. When we cross reference these individuals with those showing the highest differences in ratings between the two visits, several of these subjects came up. Rating differences of 1 or 2 points across a few dynamic and reactive items were seen. Cervical incomplete injuries have varying degrees of functional preservation below their level of injury. This could lead to inconsistent movement /compensation strategies and explain at least some of the differences in ratings.

Intra-rater reliability was also just shy of excellent (0.89). This was somewhat surprising. It also indicates that the slightly lower test re-test reliability did not solely arise from the cervical subjects performance. Further support for this line of thinking is that Palermo et al showed excellent (0.98) reliability (they called it intra-rater) across 2 visits, within 2 weeks, when using their FIST-SCI. Their cohort had a substantial number of tetraplegic subjects. Additionally, during the inter-rater reliability testing SCI sub-grouping shows a dip in reliability for the high thoracic group (0.78), instead of the cervical (0.90). Collectively, these results lead to the conclusion that differences in mFIST scores may also be attributed to our study methodology and testing environment, and it is not a direct result of variations in any given sub-group of subjects. We included a higher number of raters than any of the prior studies (6 total evaluators for the test re-test and 3 for the inter-rater) with various experience levels with the mFIST and FIST. We did not extensively train our raters prior to the start of the study. As mentioned in the methods section, this was intentional so we could see the ‘real world’ application of the mFIST. When implementing new clinical measures, evaluators often take only a small amount of training on the use of the measure. Some of the inconsistencies in mFIST ratings may reflect the slight inconsistencies both within and across the raters themselves. Indeed, if we separate subjects by raters we do see some differences; some raters had high (>0.90) reliability across their subjects while others were lower. Our results may, therefore, indicate that although the mFIST and similar measures are designed to be fairly easy to use and with little expertise, they still require that attention be paid not only to the included testing items, but also the wording, instructions and directions for users. These results may also indicate that the reliability of the FIST for SCI subjects may not be as high as previously reported in the ‘real world’.

Reliability of the mFIST was consistently excellent for lower thoracic SCI sub-groups for this entire study. Majority of the muscles of the trunk that assist with trunk control are innervated at T7 level and below. ^23–25^ With majority of these muscles intact, lower thoracic subjects have more trunk control which can lead to more reliable and consistent movements patterns. Compared to cervical and high thoracic SCI sub-groups, those with lower thoracic injuries also traditionally have a more erect sitting posture/ability. This requires less guarding and can make it easier for raters to see movement, which may make ratings more consistent.

One of the important take-home messages from this study is identifying the patterns in rating differences across the mFIST testing items. Differences between ratings were highest in dynamic testing items compared to the sitting and reactive items, across reliability testing parameters and SCI sub-groups. We initially thought this was due to the larger numbers of testing items: there are 7 different tasks in this domain compared to 4 and 3 for the sitting and reactive groups respectively. However, if we look at the frequency of rating differences > 1 across all the individual items and ratings, we continue to see that the dynamic tasks have the highest occurrence of rating differences. Rating differences in-person (test re-test) and intra-rater occurred the most (occurred ~40% of the time) with ‘picking up an object from behind’ ‘picking object from the floor’, and ‘lateral reaching’ tasks (data not shown). For interrater differences, picking up an object from the floor’, ‘forward reaching’ and ‘lateral reaching’ also occurred frequently (~40%), along with ‘posterior scooting’ coming in a close 4^th^ (data not shown). Informal feedback from raters suggested that arm placement during the ‘lateral reaching’ task and ‘picking the object from the floor’ was not detailed enough on the mFIST scoring form: raters were varied in where they allowed the subjects to place them during testing and whether that should be reflected in their scores (e.g. in the event that the untested arm was used for counter balancing). Raters were also inconsistent with how much they allowed counter balancing across dynamic and reactive tasks. Another area of inconsistency was how far a subject had to twist their trunk before grabbing the water bottle to receive a full score, and whether sitting posture should be considered in the scoring for the leaning tasks. It is reasonable to assume that these unclarified areas lead to some indecision and scoring differences.

Comments/suggestions for improvement were to either make the instructions/allowances more explicit on the mFIST form or perhaps change the rating scale. This was mostly related to the dynamic testing items where rating differences were between ‘independent’, ‘verbal cues’, and ‘upper extremity support’ making 1 and occasionally 2-point differences. This could explain why Palermo’s study had higher reliability. Their FIST-SCI measure removed ‘verbal cues’ from the rating scale. The requirements for full range of motion and/or trunk postures were also not as strict. Streamlining the scoring in these ways could make interpretations easier and thus improve inter- and intrarater reliability. It will be interesting to see, however, how the FIST-SCI and either this or another version of the mFIST perform during sensitivity studies when tracking acute or sub-acute SCI where small levels of change may be occurring and when patients are still trying to adopt movement strategies.

## Limitations

Although attempts were made to adopt a ‘real world’ experimental design, similar to all research, this study had limitations. Four out of the six in-person raters recruited their own patients for the study, which may have led to bias as the raters had familiarity of the participants prior to the tests. The mFIST was also administered in a busy clinical setting with time constraints and reduced test site availability, which could have negative implications on the amount of time for instructions or judgements on scores.

This study was exclusively administered to Veterans with SCI who receive care in the VA. It has been shown that Veterans are more likely to be older, have longer injury durations, and are often diagnosed with more secondary conditions compared to civilian counterparts.^27–31^ This could impact their ability to fully perform certain testing items, as instructed. We did not document existing secondary comorbidities, nor the presence or absence of interfering factors like pain at each testing visit. It is also impossible to know how the video scoring affected ratings. We adopted a similar protocol to Gorman et al for the inter-rater reliability because our study design required in-clinic testing. It is nearly impossible to get 3 clinical PT/DPTs at the same time to score every subject when they have their own patients.

Our sample size of 42 (with comparable SCI demographics and injury characteristics comparable to US stats) is similar to Palermo’s sample, but larger than Abou’s. As mentioned in Palermo’s study this size is large compared to other psychometric studies on trunk outcomes. ^11,14,32–34^ However, the most ideal situation that would significantly boost generalizability would be samples sizes around the 100’s and across various sites. Indeed, the smaller sample size of this study may have impacted confidence intervals and coefficients during intra-class correlations computations where a few subjects could easily skew results. This is especially true when sub-grouping SCI levels, where numbers became less than 20 per group.

## Conclusion

The mFIST has good to excellent reliability when used in a routine, busy, clinical setting with multiple raters. Reliability was consistently excellent in lower thoracic injuries. However, larger amounts of variability in scoring were seen during dynamic tasks and in those with cervical injuries. These results indicate that the mFIST might need further adjustments before being widely distributed. Determining the feasibility, validity and sensitivity of the mFIST during acute and sub-acute phases is also needed.

## Data Availability

Data produced in this study is available upon reasonable request and after approved by the Long Beach VA.

## Acknowledgements

We would like to thank Joan Judy, PTA, who was the coordinator for this study and helped with the example photos. We would also like to thank Quanisha Burnett for being a rater in this study, an author that may be added at a later date.

## Author contributions

MNH and MJ conceived this study. MNH obtained funding for the study, designed the protocol and oversaw the IRB study management. MJ, MH and VCA participated in modifying the FIST, with PK beta testing. PK, MJ, MNH, VCA and acknowledged PTs above performed all ratings with mFIST. MNH performed analyses. NV and VCA wrote portions of the manuscript, tables and/or figures with MNH oversight.

