## Supplemental Table 1 for "A Modified Version of the Function In Sitting Test (mFIST): Development and initial Reliability of the mFIST in Outpatient Clinic in Veterans with Spinal Cord Injury"

**Supplemental Table 1**: Types of Modifications to FIST

| Original FIST | | Modified FIST† |
|---|---|---|
| Anterior nudge<br>Lateral nudge<br>Posterior nudge | | No change<br>No change<br>No change |
| Static Sitting : eye open<br>Static sitting: eye closed<br>Sitting: head rotation (L/R)<br>Sitting: lift leg | | Similar -- but with palms facing upward<br>No change<br>Allowed to lift either leg of their choice. *Allowed to lift leg using upper extremity without downgrading their score* |
| Pick up object from behind<br>- Object at midline, hands breadth posterior to sacrum<br>- Have subject rotate trunk and move arm while balancing. Not allowed to blindly reach or use internal shoulder rotation<br><br>Pick up object from floor<br>- Object placed between metatarsal phalangeal joints<br>- Allowed to use either hand for retrieval<br><br>Posterior scooting<br>Anterior scooting<br>Lateral scooting<br>Lateral reach<br>- Demonstrate task first<br>- Lift dominant/stronger arm to shoulder height<br>- Reach to get weight off opposite bottom (does not have to lift off surface) | | Slight alteration<br>- Object changed to *empty water bottle* (easier to grasp)<br>- Object placed at midline, hands breadth posterior to sacrum<br>- *Touching but not grasping (due to hand function) was allowed with no penalty.*<br>- *Subject instructed to rotate trunk, if possible, to retrieve. Rotation included opposite shoulder moving forward*<br>- Allowed to pick stronger side<br>- *Allowed to do seated pivot without downgrading of score*<br>Similar but object changed to water bottle<br>- *Instructed to not use opposite arm for bracing*<br>- *Allowed to touch but not fully grasp water bottle*<br>- Allowed to pick arm for retrieval<br>No change<br>No change<br>No change<br>Slight alterations<br>- Must demonstrate example of full score<br>- Lift dominant arm to shoulder height (if possible) reach until opposite hip lifts off mat and return<br>- *Scoring for full (4) score requires maintaining upright trunk, trunk shortening, clearance of ischial tuberosity and return*<br>- *Score of 3 if performs without UE assistance but 1 or more of the requirements for full score* |

| | | |
|---|---|---|
| | | *are not met, or if subject needs multiple attempts to complete*<br>- *Opposite hand on lap for instructions. But no penalty of counter balancing with opposite arm in instructions*<br>- *No penalty if arm did not remain fully abducted during the task* |
| Forward reach<br>- Opposite arm on lap<br>- Reaching arm start 90 forward flexion<br>- Subject must go through full range of motion or until abdomen hits anterior thigh<br>- Reach with stronger/or dominant arm | | Slight changes<br>- Opposite arm on lap, *palm up*<br>- *Demonstrate first with proper form – arm at 90, head gradually facing toward floor during mid to late reach, with return not using opposite arm.*<br>- Stronger/ dominant side used<br>- *Full score (4) includes movement through full ROM or when abdomen hits anterior thighs and returns to upright sitting without bracing or use of opposite arm*<br>- *No penalty if arm was not able to be in full 90 flexion* |

[illegible]
