## Supplemental Table 2 for "A Modified Version of the Function In Sitting Test (mFIST): Development and initial Reliability of the mFIST in Outpatient Clinic in Veterans with Spinal Cord Injury"

**Supplemental Table 2: mFIST scoring sheet**

Date administered: _ _/ _ _/ _ _ _ _

Participant Instructions:

- *"Try to do each task by yourself and ask for help if you need it."*
- *"Try to do each task to the fullest extent of your ability and come back to starting position."*
- *"You may use your hands if needed for balance but try to do each task without your hands first."*
- *"Use your stronger side for one-sided tasks."*

| **Starting Position:** | **Scoring Key:** |
|---|---|
| ½ femur on surface<br>Hips & knees at 90 degrees<br>□ Check here if used step/stool for positioning or foot support | 4= Independent<br>3= Verbal cues/increased time<br>2= Upper extremity bracing/support for balance<br>1= Needs assistance to complete task<br>*document level of assist needed (mid/mod/max)<br>0= Dependent (unable to complete task, even with physical assist) |

### mFIST Test

| | | Score |
|---|---|---|
| **Randomly administer each nudge once.** Use two fingers to nudge. | **Anterior nudge: to superior sternum** | |
| | **Posterior nudge: between scapular spines** | |
| | **Lateral nudge: to dominant side at acromion** | |

| | Score |
|---|---|
| **Static sitting: 30 seconds**<br>"*Sit with your hands in your lap*."<br>Hands may be clasped together or palms facing upward. | |
| **Sitting, eyes closed: 30 seconds**<br>"*Close your eyes and remain sitting with your hands in your lap, palms up*." | |
| **Sitting, head rotation: left and right**<br>"*Sit upright and look forward. Now, look right and hold for 2 sec. Now, look left and hold for 2 sec. Return your head to center and look forward*." | |
| **Sitting, lift thigh: foot must clear ground 1 inch** **Circle leg lifted: R / L**<br>"*Lift one thigh so that your foot comes 1 inch off the ground and hold for 2 sec*."<br>Allowed to use upper extremities to lift leg without downgrading score. | |

| | Score |
|---|---|
| **Pick up object from behind: object (empty water bottle) at midline, hand's breadth posterior to sacrum**<br>"*Turn around and pick up the water bottle I've placed behind you*."<br>Seated pivot allowed without downgrading score. | |
| **Pick up object from floor: between feet at 1st MTP joint**<br>"*Pick up the water bottle from the floor and then return to sitting position*." | |

Place yardstick or 2" wide tape on mat behind and alongside patient.

| | | Score |
|---|---|---|
| **Posterior scooting: move backwards 2 inches** | *"Scoot backwards 2 inches."* | |
| **Anterior scooting: move forward 2 inches** | *"Scoot forward 2 inches towards the edge of the bed."* | |
| **Lateral scooting: move to dominant side 2 inches** | *"Scoot to the side 2 inches."* | |

| |
|---|
| **Lateral Reach: use dominant arm, must clear opposite ischial tuberosity** **Circle arm used: R / L**<br>Demonstrate task first.<br>*"Keep your feet on the floor. Lift your dominant arm out to the side at shoulder height. Reach to the side until your opposite hip lifts off the mat and come back to seated."*<br>Full score includes: maintaining upright upper trunk, abducted upper extremity, contralateral trunk shortening, clearance of contralateral ischial tuberosity, and return to sitting. **Length of reach: ______** |
| **Forward Reach: use dominant arm, full available ROM**<br>Demonstrate task first and perform movement passively to assess patient's full ROM.<br>*"Reach with your dominant arm as far as you can while staying balanced. Keep your other hand, palm up remaining in your lap."*<br>Full score includes: movement through full available ROM or when abdomen contacts anterior thighs and come back to sitting.<br>**Circle arm: R / L Length acromion to wrist:______Length of reach:______Reach/Arm length:_____%** |

| |
|---|
| **Time to administer test: _____________minutes** **Total Score** |

**Comments:** *Document if patient had any of the following: pain interference, presentation of central cord syndrome, limitations in ROM, or needs for assistance.*
